# Analysis of the correlation between anti-MDA5 antibody and the severity of COVID-19: a retrospective study

**DOI:** 10.1101/2020.07.29.20164780

**Authors:** Changzheng Liu, Qian Wang, Yeming Wang, Geng Wang, Linghang Wang, Hong Chen, Tao Jiao, Chaojun Hu, Xiaobo Lei, Li Guo, Lili Ren, Mengtao Li, Xiaofeng Zeng, Dingyu Zhang, Bin Cao, Jianwei Wang

**Author notes:** Corresponding authors: Jianwei Wang, Bin Cao Prof. Jianwei Wang., NHC Key Laboratory of Systems Biology of Pathogens and Christophe Merieux Laboratory, Institute of Pathogen Biology, Chinese Academy of Medical Sciences & Peking Union Medical College, Beijing 100730, China, Address: No. 9 Dong Dan San Tiao, Dong Cheng District, Beijing, 100730, China. Prof. Bin Cao, Department of Pulmonary and Critical Care Medicine, China-Japan Friendship Hospital; National Clinical Research Center of Respiratory Diseases; Capital Medical University; Tsinghua University-Peking University Joint Center for Life Sciences. Address: No 2, East Yinghua Road, Chaoyang District, Beijing, China. 100029.

## Abstract

**OBJECTIVE:** To identify the anti-melanoma differentiation-associated gene 5 (MDA5) antibody (Ab) in coronavirus disease 2019 (COVID-19) and its relationship with the severity and clinical outcomes of COVID-19.

**DESIGN:** Retrospective cohort study.

**SETTING:** Three hospitals in China.

**PARTICIPANTS:** 274 adult inpatients diagnosed with COVID-19 according to the Protocol for Prevention and Control of COVID-19 (Edition 7) of China and confirmed by severe acute respiratory syndrome coronavirus 2 (SARS-CoV2) RNA testing, were included from three hospitals from Wuhan, Harbin and Beijing, China from 1 December 2019 to 19 April 2020. The Biobank of Myositis Registry Department of Rheumatology, Peking Union Medical College Hospital, provided the plasma of five patients with anti-MDA5 Ab-related dermatomyositis as positive control group. Demographic, clinical and laboratory data were collected from medical records. The anti-MDA5 Ab was determined by an ELISA assay and was verified by immunoblotting analysis.

**MAIN OUTCOME MEASURES:** In hospital death of all cause.

**RESULTS:** The positive rate of anti-MDA5 Ab in patients with COVID-19 was 48.2% (132/274) and the anti-MDA5 Ab positive patients tended to represent with severe disease (88.6% *vs* 66.9%, *P*<0.0001). The titer of anti-MDA5 Ab was significantly elevated in the non-survivals (5.95±5.16 *vs* 8.22±6.64, *P*=0.030) and the positive rate was also higher than that in the survivals (23.5% *vs* 12.0%, *P*=0.012). Regarding to severe COVID-19 patients, we found that high titer of anti-MDA5 Ab (≥10.0 U/mL) was more prevalent in the non-survivals (31.2% *vs* 14.0%, *P*=0.006). Moreover, early profiling of anti-MDA5 Ab could distinguish severe patients from those with non-severe ones.

**CONCLUSION:** Anti-MDA5 Ab was prevalent in the COVID-19 patients and high titer of this antibody is correlated with severe disease and unfavorable outcomes. Early screening and serially monitoring of anti-MDA5 Ab titer have the potential to predict the disease progression of COVID-19.

## Introduction

Coronavirus Disease 2019 (COVID-19), caused by highly contagious severe acute respiratory syndrome coronavirus 2 (SARS-CoV2), has become a pandemic involving more than 12 million cases globally by July 2020.^1^ The average mortality is estimated to be 1%,^2^ but can raise up to 62% in critically ill patients, mostly due to acute respiratory distress syndrome (ARDS).^3^ Therefore, reducing mortality of severe COVID-19 patients has become an urgent task in this battle. Unfortunately, few antiviral agents have been proved to be effective enough to treat the disease, and whether to use corticosteroids and other immunomodulatory drugs remains controversial.^4^

Accumulating evidence has demonstrated that high prevalence of antinuclear antibodies (35.6%) and lupus anticoagulant (46.6%), along with antineutrophil cytoplasmic antibodies (6.6%) and anti-Ro antibody (4.4%), were also identified in hospitalized patients with COVID-19.^5^ Thus, hypothesis that SARS-CoV2 might trigger autoimmune and/or autoinflammatory aberrance in genetically predisposed subjects has been raised.^6^ Interestingly, striking similarities have been noted between multifaceted features of COVID-19 and a rare autoimmune disease, the anti-melanoma-differentiation-associated gene 5 (MDA5) antibody (Ab)-related dermatomyositis (DM).^7 8^ Both diseases can involve the lungs, skin^9 10^ and muscles.^11^ The initial radiological features of lung in Anti-MDA5 Ab-related DM patients are mainly subpleural ground-glass opacities or mixed with consolidation and signs of ARDS, which resemble severe and critical COVID-19.^12,13^ Besides, similar skin eruptions have been reported in both diseases.^10^ Furthermore, serum cytokine profiles are also similar in these two conditions, as serum levels of ferritin, IL-6, IL-8, and IL-10 usually were elevated in patients with RP-ILD secondary to Anti-MDA5 Ab-related DM.^14^ The similarity of these two diseases implies a shared underlying autoinflammatory/autoimmune mechanisms. To date, there is no report on whether anti-MDA5 Ab also exists in patients with COVID-19. It is well-known that MDA5 is a crucial cytoplasmic sensor for viral RNA and its expression is induced by RNA viruses. This activates the expression of antiviral type I and III interferons (IFNs) with inflammatory cytokines. Correspondingly, IFN signaling can induce the expression of MDA5^15^. SARS-CoV2 infection has been reported to trigger the expression of MDA5.^16 17^ In addition, MDA5 is involved in pathogenesis of several autoimmune disorders as well,^15^ such as systemic lupus erythematosus^18 19^, multiple sclerosis^20^, and even type 1 diabetes^21 22^. Nevertheless, it remains unclear whether the anti-MDA5 Ab plays a role in the pathophysiology of COVID-19 or whether it correlates with the disease severity. Some researchers have called for screening the anti-MDA5 Ab in severe COVID-19 patients.^7 8^ In this study, we investigate the presence of anti-MDA5 Ab in patients with SARS-CoV2 infection and address its correlation with the clinical severity and outcomes of COVID-19.

## Methods

### Study design and population

This retrospective study included three cohorts of adult patients (≥18 years old) from Jinyintan Hospital (Wuhan, China), Beijing Ditan Hospital (Beijing, China), and Heilongjiang Infectious Disease Hospital (Harbin, China), who were hospitalized from Dec 1, 2019 to Apr 19, 2020. All patients who were diagnosed with COVID-19 according to the Protocol for Prevention and Control of COVID-19 (Edition 7) promulgated by National Health Commission of China.^23^ All patients with COVID-19 were tested positive for SARS-CoV2 by use of quantitative, real-time PCR technology on samples from the upper airway specimens (pharyngeal or nasal swabs, nasopharyngeal secretions). The plasma of patients with COVID-19 were collected within 24 hours after admission, and stored in -80°C. The plasma of five patients with anti-MDA5 Ab-related DM were provided by the Biobank of Myositis Registry of Department of Rheumatology, Peking Union Medical College Hospital. All of the 5 DM patients were diagnosed based on the criteria of Bohan and Peter.^24 25^

The study was approved by the Research Ethics Committee of the participating hospitals and the ethical board of the Institute of Pathogen Biology, Chinese Academy of Medical Sciences. The requirement for informed consent was waived by the Ethics Commission of the designated hospitals for emerging infectious diseases as described previously^12^.

### Data collection

We extracted demographic, clinical, laboratory, treatment, and outcome data from medical and nursing records using standardized data collection forms (a revised version of case record form for severe acute respiratory infection shared by WHO and the International Severe Acute Respiratory and Emerging Infection Consortium). All data were checked by two investigators and a third researcher adjudicated any difference in interpretation between the two primary reviewers.

According to the clinical classification of COVID-19 by the Protocol for Prevention and Control of cases of COVID-19 (Edition 7),^23^ we divided the patients into two groups on hospitalization: (1) Severe group, the patients fulfilled the diagnostic criteria of severe and critical cases, who meets any one of follows: i) Respiratory rate > 30/min, ii) Pulse oxygen saturation < 93%, iii) Oxygenation index < 300 mmHg, or iv) respiratory failure or other organ dysfunction requiring transmission to intensive care unit. (2) Non-severe group, the patients’ severity was mild or moderate that didn’t meet the above criteria.

### Western Blots

293T cell line was obtained from the American Type Culture Collection (ATCC) and grown in DMEM with 10% FBS (Hyclone) at 37°C in 5% CO_2_ cell culture incubator. This cell line was tested 1 month before the experiment by methods of morphology check by microscopy, growth curve analysis, and mycoplasma detection according to the ATCC cell line verification test recommendations. The transfection of the plasmid expressing human MDA5 cDNA was performed using Lipofectamine 2000 transfection reagent (Invitrogen) according to the manufacturer’s instruction. Western blotting of proteins was performed as described previously^26^. The antibodies used included those against Flag and MDA5 were purchased from Sigma-Aldrich Co. and the antibody against β-actin were obtained from Abcam Co.

### ELISA

IgG against MDA5 were detected in plasma samples using Anti-MDA5 Ab ELISA Kits (Medical & Biological Laboratories Co.), according to the manufacturer’s instructions. Briefly, during plasma samples incubation, anti-MDA5 Ab in the plasma reacts with the immobilized human MDA5 antigen coated in the Microwells before. The test result is determined photometrically by measuring the absorbance (wave length: 450nm) and plotting the results performed on microplate reader (SpectraMax M5, MD, USA). The formula used for the unit value calculation is: Unit value (U/ml) =(A450 < Sample > - A450 < Calibrator 1> / (A450 < Calibrator 2 > -A450 < Calibrator1 >)×100, which refers to the manufacturer’s instructions. The unit value ≥ 5.0 U/mL is considered positive and the unit value ≥ 10.0 U/mL is defined as high titer of anti-MDA5 Ab.

### Statistical analysis

For the detection of anti-MDA5 Ab, each experiment was repeated 3 times. Unpaired, two-sided Mann-Whitney *U*-test was performed to compare two groups unless otherwise indicated (X2 test). For the clinical analysis of anti-MDA5 Ab, descriptive statistics (percentages, means, standard deviations [SDs], medians, interquartile [IQR]) were provided for describe baseline demographic and clinical characteristics. The comparison of demographic, clinical, laboratory characteristics and outcomes across anti-MDA5 Ab positive/negative and survival/non-survival subgroups was performed by the Chi-squared tests or analysis of variance as appropriate. All statistical analyses were performed using SPSS 16.0 software (SPSS Inc., Chicago, IL, USA). *P*-values <0.05 were considered statistically significant.

## Results

### Patients description

A total of 274 patients were included in this study (Figure 1a; Table 1). Of these patients, 230 cases from Wuhan Jinyintan hospital, the time admitted to hospital is from Dec 1, 2019 to Jan 27, 2020. 24 cases from Harbin infectious disease hospital, the time admitted to hospital is from Apr 2 to Apr 19. 20 cases from Beijing Ditan hospital, the time admitted to hospital is from Jan 20, 2020 to Jan 26, 2020. The median age was 56 years (IQR, 45-65 years), and 159 (58.0%) patients were male. The average disease course from onset of symptoms to discharge was 22.8±9.6 days. According to the definition of disease severity, 212 (77.4%) patients were classified as severe disease. Nearly half of the patients (n=119, 43.4%) had underlying chronic diseases, including hypertension, coronary arterial disease, chronic lung disease, and diabetes mellitus. On admission, 43 (15.7%) patients were complicated with respiratory failure, shock or other organ dysfunctions. 31 (11.3%) were transferred to intensive care unit during their hospital stay, and 48 (17.5%) patients died.

**Figure 1.** **Anti-MDA5 Ab is determined in patients with COVID-19. a**, Overview of the cohort in this study, including healthy donors (n=50) and patients with COVID-19 (n=274). Of these patients, 62 non-severe COVID-19 patients included mild and moderate clinical performance, which was defined by the symptoms with or without mild lung change. Severe disease status (n=212): Clinical symptoms with severe lung change (Lesions progression > 50%), organ dysfunction, respiratory failure, shock, and intensive care unit (ICU) admission, and decease (n=48). **b**, The titer of anti-MDA5 Ab was increased in patients with COVID-19. The plasma samples from healthy donors served as normal control (Normal). **c**, Graph of positive rate of anti-MDA5 Ab in COVID-19 was higher than that in normal control (132/174, 48.18%). The numbers of normal control and COVID-19 patients are indicated underneath. *P* values were determined by using unpaired, two-sided Mann-Whitney *U*-test and X2 test. *P*<0.0001, ****. **d, e, f, and g**, MDA5 overexpression (OE) was achieved in 293-T cells and immunoblots were performed with Anti-MDA5 Ab, Anti-FLAG Ab (d), plasma form DM (e), and plasma from COVID-19 patients (f and g). (β-actin is used as a loading control and the unit values from ELISA of each COVID-19 plasma samples are shown underneath.

**Table 1.**
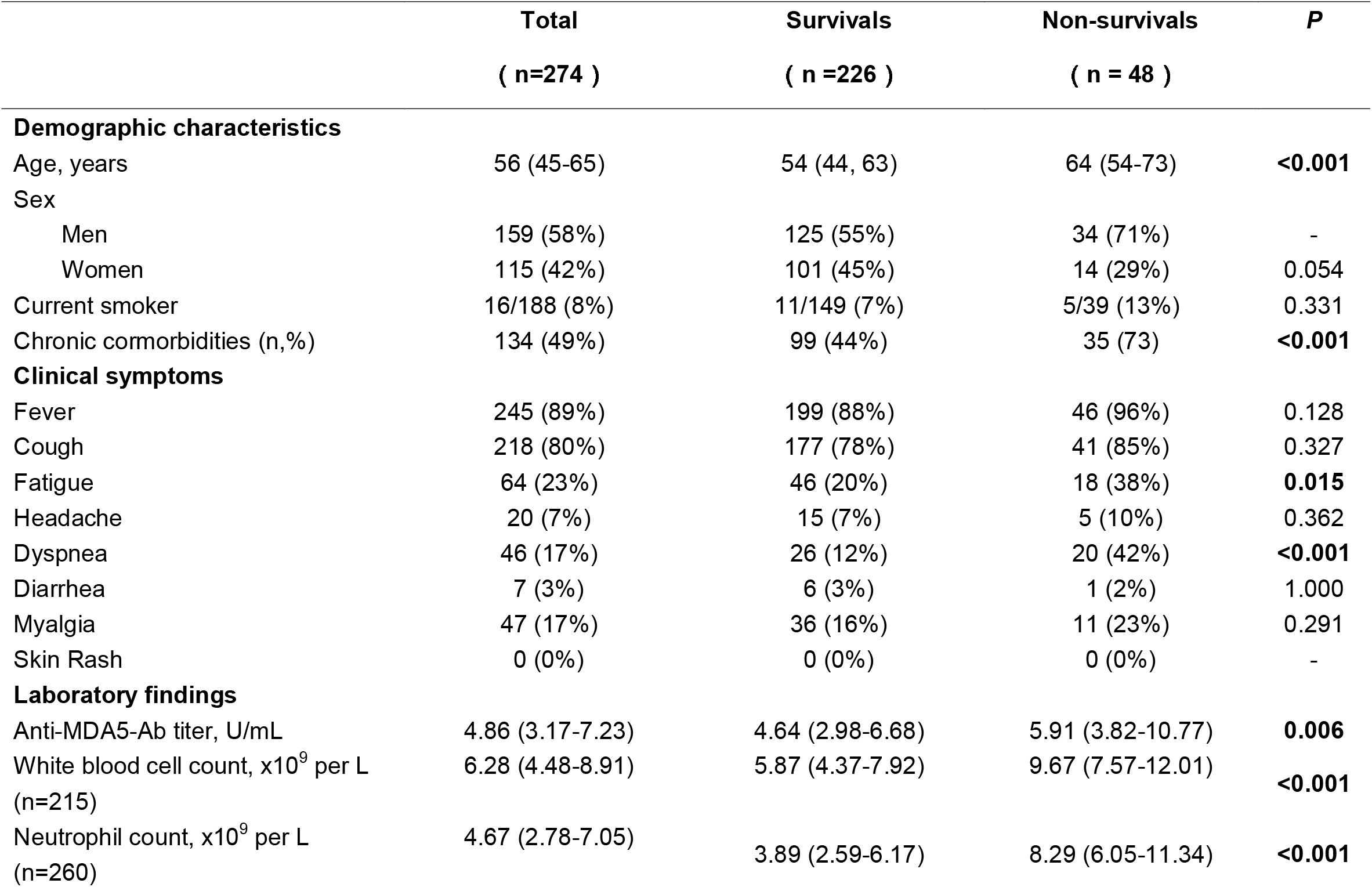
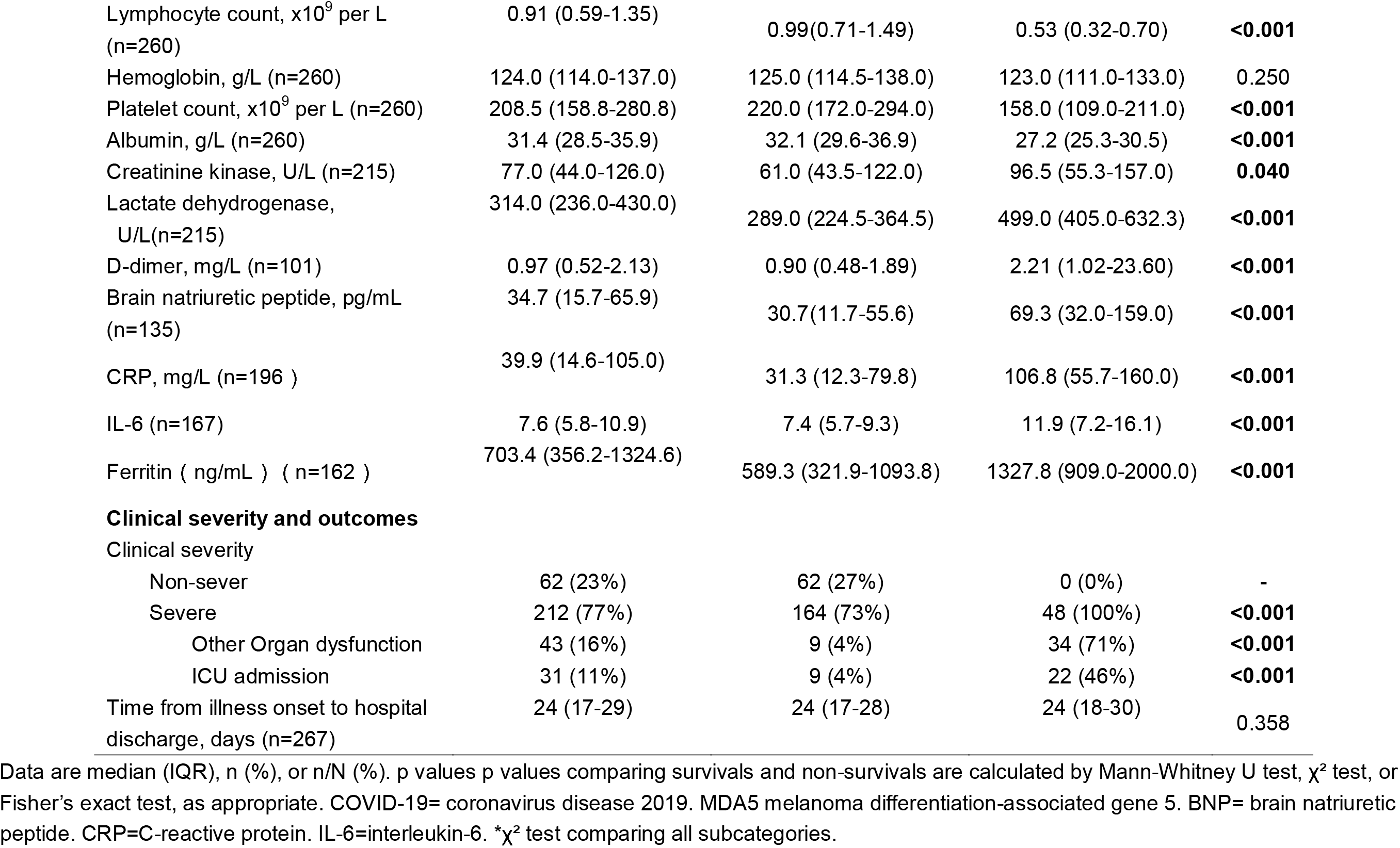
Demographic, clinical, laboratory findings, and outcomes of patients with COVID-19.

|  | **Total** | **Survivals** | **Non-survivals** | **P** |
| --- | --- | --- | --- | --- |
|  | **( n=274 )** | **( n =226 )** | **( n = 48 )** |  |
| **Demographic characteristics** |  |  |  |  |
| Age, years | 56 (45-65) | 54 (44, 63) | 64 (54-73) | **<0.001** |
| Sex |  |  |  |  |
| Men | 159 (58%) | 125 (55%) | 34 (71%) | - |
| Women | 115 (42%) | 101 (45%) | 14 (29%) | 0.054 |
| Current smoker | 16/188 (8%) | 11/149 (7%) | 5/39 (13%) | 0.331 |
| Chronic cormorbidities (n,%) | 134 (49%) | 99 (44%) | 35 (73) | **<0.001** |
| **Clinical symptoms** |  |  |  |  |
| Fever | 245 (89%) | 199 (88%) | 46 (96%) | 0.128 |
| Cough | 218 (80%) | 177 (78%) | 41 (85%) | 0.327 |
| Fatigue | 64 (23%) | 46 (20%) | 18 (38%) | **0.015** |
| Headache | 20 (7%) | 15 (7%) | 5 (10%) | 0.362 |
| Dyspnea | 46 (17%) | 26 (12%) | 20 (42%) | **<0.001** |
| Diarrhea | 7 (3%) | 6 (3%) | 1 (2%) | 1.000 |
| Myalgia | 47 (17%) | 36 (16%) | 11 (23%) | 0.291 |
| Skin Rash | 0 (0%) | 0 (0%) | 0 (0%) | - |
| **Laboratory findings** |  |  |  |  |
| Anti-MDA5-Ab titer, U/mL | 4.86 (3.17-7.23) | 4.64 (2.98-6.68) | 5.91 (3.82-10.77) | **0.006** |
| White blood cell count, x10<sup>9</sup> per L<br>(n=215) | 6.28 (4.48-8.91) | 5.87 (4.37-7.92) | 9.67 (7.57-12.01) | **<0.001** |
| Neutrophil count, x10<sup>9</sup> per L<br>(n=260) | 4.67 (2.78-7.05) | 3.89 (2.59-6.17) | 8.29 (6.05-11.34) | **<0.001** |
| Lymphocyte count, x10<sup>9</sup> per L (n=260) | 0.91 (0.59-1.35) | 0.99(0.71-1.49) | 0.53 (0.32-0.70) | **<0.001** |
| Hemoglobin, g/L (n=260) | 124.0 (114.0-137.0) | 125.0 (114.5-138.0) | 123.0 (111.0-133.0) | 0.250 |
| Platelet count, x10<sup>9</sup> per L (n=260) | 208.5 (158.8-280.8) | 220.0 (172.0-294.0) | 158.0 (109.0-211.0) | **<0.001** |
| Albumin, g/L (n=260) | 31.4 (28.5-35.9) | 32.1 (29.6-36.9) | 27.2 (25.3-30.5) | **<0.001** |
| Creatinine kinase, U/L (n=215) | 77.0 (44.0-126.0) | 61.0 (43.5-122.0) | 96.5 (55.3-157.0) | **0.040** |
| Lactate dehydrogenase, U/L(n=215) | 314.0 (236.0-430.0) | 289.0 (224.5-364.5) | 499.0 (405.0-632.3) | **<0.001** |
| D-dimer, mg/L (n=101) | 0.97 (0.52-2.13) | 0.90 (0.48-1.89) | 2.21 (1.02-23.60) | **<0.001** |
| Brain natriuretic peptide, pg/mL (n=135) | 34.7 (15.7-65.9) | 30.7(11.7-55.6) | 69.3 (32.0-159.0) | **<0.001** |
| CRP, mg/L (n=196 ) | 39.9 (14.6-105.0) | 31.3 (12.3-79.8) | 106.8 (55.7-160.0) | **<0.001** |
| IL-6 (n=167) | 7.6 (5.8-10.9) | 7.4 (5.7-9.3) | 11.9 (7.2-16.1) | **<0.001** |
| Ferritin ( ng/mL ) ( n=162 ) | 703.4 (356.2-1324.6) | 589.3 (321.9-1093.8) | 1327.8 (909.0-2000.0) | **<0.001** |
| **Clinical severity and outcomes** |  |  |  |  |
| Clinical severity |  |  |  |  |
| Non-sever | 62 (23%) | 62 (27%) | 0 (0%) | - |
| Severe | 212 (77%) | 164 (73%) | 48 (100%) | **<0.001** |
| Other Organ dysfunction | 43 (16%) | 9 (4%) | 34 (71%) | **<0.001** |
| ICU admission | 31 (11%) | 9 (4%) | 22 (46%) | **<0.001** |
| Time from illness onset to hospital discharge, days (n=267) | 24 (17-29) | 24 (17-28) | 24 (18-30) | 0.358 |

Data are median (IQR), n (%), or n/N (%). p values p values comparing survivals and non-survivals are calculated by Mann-Whitney U test, $\chi^2$ test, or Fisher's exact test, as appropriate. COVID-19= coronavirus disease 2019. MDA5 melanoma differentiation-associated gene 5. BNP= brain natriuretic
peptide. CRP=C-reactive protein. IL-6=interleukin-6. \* $\chi^2$ test comparing all subcategories.

### Anti-MDA5 Ab is identified in the plasma of patients with COVID-19

To determine the presence of anti-MDA5 Ab in COVID-19 patients, ELISA analysis was employed to test the plasma collected from COVID-19 patients. We found that the titer of anti-MDA5 Ab is increased in the plasma of COVID-19 patients as compared to normal controls (1.85±0.67 *vs* 6.60±5.50, *P*<0.0001) (Figure 1b). The plasma from five patients of anti-MDA5 Ab-related DM were used as positive controls (Supplementary Figure 1). Based on the cut-off value (5.0 U/mL) of the anti-MDA5 Ab ELISA Kit, we noticed that the positive rate of anti-MDA5 Ab was also higher in COVID-19 patients than that in normal controls (P<0.0001) (Figure 1c). These data were further validated by immunoblots in selected COVID-19 plasma samples. To this aim, we firstly performed MDA5 overexpression in 293T cells as shown by Western blotting analysis (Figure 1d). The plasma of anti-MDA5 Ab-related DM patients included in the ELISA were also confirmed (Figure 1e). Next, a total of 17 plasma samples of COVID-19 were conducted Western blotting analysis, which included five non-severe and 12 severe COVID-19 patients. The data showed that the anti-MDA5 Ab were detected in these examined samples as well (Figures 1f and 1g).

Altogether, these data indicate that SARS-CoV2 infection leads to an increased anti-MDA5 Ab titer in with COVID-19 patients.

### COVID-19 patients with positive anti-MDA5 Ab tend to exhibit severe disease

Anti-MDA5 Ab is first identified in DM and correlated with the status of this disease, which promoted us to further investigate whether the titer of anti-MDA5 Ab was related to the clinical severity of patients with COVID-19. To this end, 274 COVID-19 patients were stratified into two groups based on the cutoff value, that is, the anti-MDA5 Ab negative (<5.0 U/mL) and positive (≥5.0 U/mL). We found that the percentage of severe COVID-19 patients was much higher in anti-MDA5 Ab positive group than that in negative group (88.6% *vs* 66.9%, *P*<0.0001) (Figure 2a). The survival rate of anti-MDA5 Ab positive patients with COVID-19 is much lower compared with the negative group (76.5% *vs* 88.0%, *P*=0.012) (Figure 2b). As expected, the COVID-19 patients with positive anti-MDA5 Ab tended to have much longer disease course at discharge and develop respiratory failure, shock and other organ dysfunction (Figures 2c and 2d). These data indicated that COVID-19 patients with increased anti-MDA5 Ab titer exhibited severe clinical performance. Next, a univariate analysis was employed to investigate the correlation between anti-MDA5 Ab and other COVID-19 prognostic factors (Table 2). We found that the titer of anti-MDA5 Ab was positively correlated with the age of COVID-19 patients (Figure 2f). We also noticed that COVID-19 patients with positive anti-MDA5 Ab depicted decreased lymphocytes, increased neutrophils (Figures 2f and 2g). The levels of albumin were found to decrease in anti-MDA5 Ab positive patients compared with the negative (Figure 2h). The ratio of neutrophils versus lymphocytes (NLR) or CRP versus albumin (CAR) was much higher in anti-MDA5 Ab positive samples than that in the negative, indicating much severer inflammatory damage (Figures 2j and 2k). No significant difference was observed in Creatine Kinase (CK), lactate dehydrogenase (LDH), ferritin, and CRP (Table 2).

**Figure 2.** **COVID-19 patients with positive anti-MDA5 Ab exhibit severe clinical performance. a**, Comparison of the percentage of COVID-19 patients with non-severe (mild & moderate) and severe performance in anti-MDA5 Ab negative (Anti-MDA5 Ab Neg) and anti-MDA5 Ab positive (Anti-MDA5 Ab Pos) group. **b**, The survival rate is higher in anti-MDA5 Ab Neg group than that in anti-MDA5 Ab Pos group. **c**, Comparison of the total disease course of COVID-19 patients with positive and negative anti-MDA5 Ab. **d**, Comparison of the percentage of COVID-19 patients with organ dysfunction in anti-MDA5 Ab Neg and anti-MDA5 Ab Pos group. **e**, Comparison of the age of COVID-19 patients in anti-MDA5 Ab Neg and anti-MDA5 Ab Pos group. **f**, The number of lymphocytes is decreased in COVID-19 patients with positive anti-MDA5 Ab as compared to that in anti-MDA5 Ab negative group. **g**, The number of neutrophils is increased in COVID-19 patients with positive anti-MDA5 Ab as compared to that in anti-MDA5 Ab negative group. **h**, The levels of albumin are decreased in COVID-19 patients with positive anti-MDA5 Ab compared with that in anti-MDA5 Ab negative group. **i and j**, The ratios of NLR (i) and CAR (j) are increased in anti-MDA5 Ab Pos patients with COVID-19. The numbers of COVID-19 patients in each group are indicated underneath. *P* values were determined by using unpaired, two-sided Mann-Whitney *U*-test and X2 test. *P*<0.05, *; *P*<0.01, **; *P*<0.001, ***; *P*<0.0001, ****.

**Table 2.** Demographic, clinical, laboratory findings, and outcomes of patients with COVID-19.

|  | Anti-MDA5 Ab positive<br>(n =132) | Anti-MDA5 Ab negative<br>(n = 142) | *P* |
| --- | --- | --- | --- |
| **Demographic characteristics** |  |  |  |
| Age, years | 59.0 (50.3, 68.0) | 51.00 (42.00, 63.00) | **<0.001** |
| Sex | 69 (52.3) | 90 (63.4) | 0.063 |
| Men |  |  |  |
| Women |  |  |  |
| Current smoker | 10 (9.8) | 6 (7) | 0.489 |
| Chronic cormorbidities (n,%) | 75 (56.8) | 59 (41.5) | **0.012** |
| **Clinical symptoms** |  |  |  |
| Fever | 120 (90.9) | 125 (88.0) | 0.439 |
| Cough | 117 (88.6) | 101 (71.1) | **<0.001** |
| Fatigue | 34 (25.8) | 30 (21.1) | 0.365 |
| Headache | 12 (9.1) | 8 (5.6) | 0.272 |
| Dyspnea | 25 (18.9) | 21 (14.8) | 0.358 |
| Diarrhea | 3 (2.3) | 4 (2.8) | 0.775 |
| Myalgia | 21 (15.9) | 26 (18.3) | 0.598 |
| Skin Rash |  |  |  |
| **Laboratory findings** |  |  |  |
| Anti-MDA5-Ab titer, U/mL | 7.36 (5.88, 11.04) | 3.26 (2.23, 4.19) | **<0.001** |
| White blood cell count, x10<sup>9</sup> per L<br>(n=215) | 6.9 (4.8, 9.4) | 5.81 (4.3, 8.3) | 0.079 |
| Neutrophil count, x10<sup>9</sup> per L<br>(n=260) | 5.17 (3.15, 7.90) | 3.90 (2.58, 6.43) | **0.024** |
| Lymphocyte count, x10<sup>9</sup> per L<br>(n=260) | 0.84 (0.57, 1.17) | 0.98 (0.65, 1.54) | **0.040** |
| Hemoglobin, g/L (n=260) | 123.00 (113.00, 136.25) | 128.00 (115.75, 138.00) | 0.193 |
| Platelet count, x10<sup>9</sup> per L (n=260) | 209.50 (146.00, 285.25) | 207.00 (169.50, 276.00) | 0.754 |
| Albumin, g/L (n=260) | 30.45 (27.30, 33.50) | 32.95 (29.65, 38.80) | **<0.001** |
| Creatinine kinase, U/L (n=215) | 66.00 (43.50, 118.50) | 73.50 (45.75, 158.75) | 0.474 |
| Lactate dehydrogenase, U/L<br>(n=215) | 323.00 (238.50, 447.00) | 303.50 (227.00, 419.50) | 0.410 |
| D-dimer, mg/L (n=101) | 0.85 (0.36, 1.92) | 0.41 (0.00, 1.24) | **<0.001** |
| Brain natriuretic peptide, pg/mL<br>(n=135) | 36.60 (18.60, 81.30) | 31.40 (10.00, 65.53) | 0.107 |
| CRP, mg/L (n=196 ) | 42.70 (18.08, 110.30) | 39.55 (10.30, 90.83) | 0.095 |
| IL-6 (n=167) | 7.41 (5.53, 10.09) | 8.02 (6.15, 11.39) | 0.177 |
| Ferritin (ng/mL) (n=162) | 801.31 (381.88, 1397.92) | 678.19 (320.31, 1251.58) | 0.270 |
| NLR (n=260) | 5.26 (2.97, 12.07) | 3.76 (2.24, 8.56) | **0.016** |
| PLR (n=260) | 5.17 (3.15, 7.90) | 3.90 (2.58, 6.43) | 0.193 |
| **Clinical severity and outcomes** |  |  |  |
| Clinical severity | 117 (88.6) | 95 (66.9) | **<0.001** |
| Non-sever |  |  |  |
| Severe |  |  |  |
| Other Organ dysfunction | 27 (20.5) | 16 (11.3) | **0.037** |
| ICU admission | 18 (13.6) | 13 (9.2) | 0.242 |
| Death | 31 (23.5) | 17 (12.0) | **0.012** |
| Time from illness onset to hospital discharge, days (n=267) |  |  |  |

Data are median (IQR), n (%), or n/N (%). *P*-values were calculated by Mann-Whitney U test, $\chi^2$ test, or Fisher's exact test, as appropriate. COVID-19= coronavirus disease 2019. MDA5 melanoma differentiation-associated gene 5. BNP= brain natriuretic peptide . CRP=C-reactive protein. IL-6=interleukin-6. $\chi^2$ test comparing all subcategories.

Taken together, our findings suggest that anti-MDA5 Ab is positively correlated with the clinical severity of COVID-19 patients.

### The correlation between anti-MDA5 Ab and COVID-19 outcomes

As mentioned above, COVID-19 patients with positive anti-MDA5 Ab tended to develop severe disease. The titer of anti-MDA5 Ab was higher in severe COVID-19 patients as compared to the non-severe (Figure 3a). The positive rate of this antibody was also higher in severe COVID-19 patients (Figure 3b). We also observed that the titer of anti-MDA5 Ab depicted a significant increase in COVID-19 patients suffering from the chronic comorbidities, for instance, hypertension, diabetes, and cardiovascular disease (Figure 3c). An augment of this antibody was noticed in COVID-19 patients suffering from shock, respiratory or other organ failure (Figure 3d).

**Figure 3.** **The correlation between anti-MDA5 Ab and the outcome of COVID-19 patients. a and b**, Comparison of the titer and positive rate of anti-MDA5 Ab in COVID-19 patients with non-severe and severe performance. **c and d**, The titer of anti-MDA5 Ab are increased in COVID-19 patients with chronic comorbidities (c) and organ failure (d). **e and f**, The titer (e) and positive rate (f) of anti-MDA5 Ab is higher in the deceased patients with COVID-19. **g**, Comparison of multiple varies in the survival and deceased patients with COVID-19 as shown in heatmap paragraphs. **h and i**, The titer (h) and positive rate (i) of anti-MDA5 Ab are elevated in the survival and dead patients with severe performance compared with that in the non-severe. **j**, Comparison of the percentage of patients with H-anti-MDA5 Ab (Unit value ≥ 10.0) in the survival and dead patients with severe performance. The numbers of COVID-19 patients in each group are indicated underneath. *P* values were determined by using unpaired, two-sided Mann-Whitney *U*-test and X2 test. *P*<0.05, *; *P*<0.01, **; *P*<0.0001, ****.

We next investigated whether the titer of anti-MDA5 Ab was correlated with the outcome of COVID-19 patients. To this end, a further comparison of anti-MDA5 Ab was employed in the survival COVID-19 patients and the non-survivals, which indicated that the titer of anti-MDA5 Ab was upregulated in dead patients compared with the survival (Figure 3e). Its positive rate was lower in the survivals with COVID-19 than that in the non-survivals (Figure 3f). These data suggested that anti-MDA5 Ab had the potential to serve as a prognostic factor for COVID-19. Consistent with published predictive factors for COVID-19 outcomes, we found that the levels of LDH, ferritin, and CRP were significantly decreased in the non-survivals as compared to that in the survivals, and the number of lymphocytes was also markedly reduced in the non-survivals (Figure 3g; Table 1).

We further performed a comparison of the anti-MDA5 Ab in COVID-19 patients with non-severe, severe performance and those deceased. The titer of anti-MDA5 Ab and positive rate were increased in severe and deceased patients compared with the non-severe ones (Figures 3h and 3i). Although both of the titer and positive rate of anti-MDA5 Ab depicted a moderate increase in the deceased patients as compared to the severe ones, no significant difference was observed between these two clusters (Figures 3h and 3i). In addition, we addressed the difference between the survivals and non-survivals in severe COVID-19 patients using 2-fold cut-off value based on the ELISA kit and found that the percentage of COVID-19 patients with high titer of anti-MDA5 Ab (≥ 10.0 U/mL) was elevated in the non-survivals than that in the survivals (Figure 3j).

Altogether, our data indicate that anti-MDA5 Ab is a marker for prognosis of COVID-19 patients and severe COVID-19 patients with high titer of anti-MDA5 Ab tend to have elevated mortality.

### Early profile of anti-MDA5 Ab distinguishes the prognosis of non-severe and severe COVID-19

Since the alteration of anti-MDA5 Ab titer is correlated with the activity and outcome of DM, we asked whether the change of anti-MDA5 Ab was associated with the clinical features of COVID-19. To this end, a cross-sectional analysis was employed using the titer of anti-MDA5 Ab achieved from the whole disease course. A total of 273 cases was stratified into three clusters based on the weeks following symptoms onset (WFSO) and shown as follows, WFSO-1, WFSO-2, and WFSO-3 (Figure 4a). A longitudinal profiling of anti-MDA5 Ab from 23 cases of severe COVID-19 patients was also determined at 4 sequential time-points (Figure 4a). A significant increase of the positive rate of anti-MDA5 Ab was observed in the samples from WFSO-2 and WFSO-3, as compared to WFSO-1, although no difference of the anti-MDA5 Ab titer was noticed in these three clusters (Figures 4b and 4c). These data revealed that the dynamic alteration of anti-MDA5 Ab might be various in the disease course of COVID-19 patients with diverse clinical performance. To test this idea, we compared anti-MDA5 Ab at three intervals as stated above in non-severe and severe patients, respectively. Interestingly, the titer of anti-MDA5 Ab in non-severe patients with COVID-19 was significantly increased at WFSO-2 as compared to that in WFSO-1, and then decreased at WFSO-3 (Figure 4d,). Similar result was found in the positive rate of anti-MDA5 Ab (Figure 4e). On the contrary, COVID-19 patients with severe performance exhibited high titer of anti-MDA5 Ab at the disease onset (WFSO-1) and then decreased at WFSO-2 and -3 (Figure 4f). However, no significant alteration was observed in the positive rate of this antibody at all three intervals, indicating that high titer of anti-MDA5 was preserved in the disease course of severe COVID-19 (Figure 4g).

**Figure 4.** **Overview of the anti-MDA5 Ab profile in COVID-19. a**, Overview of the cross-sectional and longitudinal analyses in COVID-19. **b and c**, A cross-sectional analysis of the anti-MDA5 Ab titer (b) and positive rate (c) is employed in 273 cases of COVID-19 patients. **d and e**, A cross-sectional analysis of the anti-MDA5 Ab titer (d) and positive rate (e) is performed in non-severe (c) COVID-19 patients. **f and g**, A cross-sectional analysis of the anti-MDA5 Ab titer (f) and positive rate (g) is performed in severe COVID-19 patients. The samples were stratified into three clusters: WFSO-1, WFSO-2, and WFSO-3. The numbers of COVID-19 patients in each cluster are indicated underneath. *P* values were determined by using unpaired, two-sided Mann-Whitney *U*-test and X2 test. *P*<0.05, *; *P*<0.01, **; *P*<0.001, ***; *P*<0.0001, ****. **h and i**, A longitudinal analysis of anti-MDA5 Ab profiling in 3 patients (h). 4 time-points were selected, which began from WFSO-1 (Patient 1, DFSO-6; Patient 2, 3, DFSO-7) (i, left panel). Longitudinal data were also plotted over time continuously according to DFSO. Regression lines are indicated using the red solid line (i, right panel). **j and k**, A longitudinal analysis of anti-MDA5 Ab profiling in 20 patients. Of them, patient 11, 12, and 14 were shown in panel j. 4 time-points were selected, which began from DWSO-2 (k, left panel). Longitudinal data were also plotted over time continuously according to DFSO. Regression lines are indicated using the red solid line (k, right panel).**l**, The positive rate of anti-MDA5 Ab was determined in 20 patients as stated above at DFSO 8-14, 13-19, 18-25, and 23-30. **m and n**, Comparison of the titer of anti-MDA5 Ab in non-severe and severe COVID-19 patients at DWSO 1, 2, and 3 (m). The cross-sectional data was also plotted according to days following symptom onset. Regression lines are indicated by the blue (non-severe) or red (severe) solid lines (n). The numbers of COVID-19 patients in each cluster are indicated underneath. *P* values were determined by using unpaired, two-sided Mann-Whitney *U*-test and X2 test. *P*<0.05, *; *P*<0.01, **; *P*<0.001, ***; *P*<0.0001, ****.

We further determined the titer of anti-MDA5 Ab in sequential samples from severe COVID-19 disease as shown in Figure 4a. The titer of anti-MDA5 Ab in patient #1, #2, and #3 depicted a similar alteration compared with that in the cross-sectional analysis (Figures 4h and 4i). Next, the titer of anti-MDA5 Ab was examined in the samples collected at the WFSO-2, that is, the days following symptoms onset (DFSO) 8-14. We found that the titer and positive rate of anti-MDA5 Ab remained substantial (Figures 4j, 4k, and 4l; Supplementary Figure 2).

Collectively, our data indicate that COVID-19 patients with high titer of anti-MDA5 Ab initially tend to develop severe disease.

## Discussion

The present study, for the first time, identified and confirmed the prevalence of anti-MDA5 Ab in patients with COVID-19 by both ELISA and Western blots. We also demonstrated that the positive rate and titer of anti-MDA5 Ab was associated with the clinical severity and outcomes of COVID-19. In severe COVID-19 patients, we found that high titer of anti-MDA5 Ab (≥10.0 U/mL) was more prevalent in non-survival patients. Moreover, early profile of anti-MDA5 Ab could distinguish severe patients from non-severe ones. Our study provides the evidence that early screening of anti-MDA5 Ab might help identify high risk population and predict the outcome of patients with COVID-19.

MDA5 is a crucial antiviral factor and has been previously reported to involve in SARS-CoV, MERS-CoV and SARS-CoV2 infections.^16, 27 28^ Interestingly, MDA5 is also involved in several autoimmune disorders such as anti-MDA5 Ab-related DM. Therefore, it is not surprising that COVID-19 and anti-MDA5 Ab-related DM share similar features of hyperinflammation and multi-systemic manifestations, especially rapid progressive interstitial lung disease that results in ARDS and death. In this study, we determined anti-MDA5 Ab in as many as 48.2% patients with COVID-19.

Our study revealed a positive correlation between the anti-MDA5 Ab and the severity of COVID-19, and high titer of anti-MDA5 Ab was associated with higher mortality in severe COVID-19 patients. Similar observation was reported in anti-MDA5 Ab-related DM patients.^29^ However, the titer of this antibody is even higher in anti-MDA5 Ab-related DM than that in COVID-19. This may indicate that high titer of anti-MDA5 Ab probably is related to an uncontrolled autoinflammation and autoimmune response to SARS-CoV2 infection in genetically predisposed hosts. Furthermore, our study also demonstrated that elder age, chronic comorbidities, lymphocytopenia, hypoalbuminemia, hyperferritinemia, increased D-dimer and CRP levels were more prevalent in COVID-19 patients with organ dysfunction and the mortality was comparatively high, which has been reported in previous studies and implies a dysregulation of inflamation. ^12 30-34^

It has been reported that the change of anti-MDA5 Ab titer correlates with disease activity and predicts treatment response and disease outcome in patients with DM and rapidly progressive interstitial lung disease. ^29^ Our data also indicated that the dynamic titer alteration of anti-MDA5 Ab clearly varied in COVID-19 patients with diverse clinical severities. In the non-severe patients, the titer of anti-MDA5 Ab is upregulated in week 2 after symptom onset and then decreased, suggesting that the IFNs-MDA5 circuit is under fine-tuning regulation and the immune homeostasis is preserved in the total process of SARS-CoV2 infection (Figures 4m and 4n). However, in the severe COVID-19 patients, the titer of anti-MDA5 Ab boosts up in the 1st week after symptom onset and subsequently remains at a high positivity although a decreased titer is observed at weeks 2, 3, and 4 (Figures 4m and 4n). These data further supported that the MDA5 signaling might be persistently over-activated in severe COVID-19 patients. These findings also suggest that early screening and serially monitoring of anti-MDA5 Ab titer has the potential to predict the disease progression of COVID-19.

Several studies have already shown effectiveness of tocilizumab (IL-6 receptor blockade)^35^, ruxolitinib (JAK inhibitor)^36^ and tacrolimus^37^ in inhibiting SARS-CoV2 replication, improving the chest tomography or facilitating clinical improvement. Recently, dexamethasone has also been reported to improve survival in severe COVID-19 patients as well.^38^ Our findings provide supportive evidence that anti-inflammation and immunosuppressive therapy might be compromising strategy for the treatment of COVID-19, especially in those with high titer of anti-MDA5 Ab.

There are several limitations in our study. Firstly, since MDA5 is validated as a general sensor for diverse RNA viruses, no evidence has addressed whether anti-MDA5 Ab is present in the infection of other RNA viruses, for instance, influenza virus, enterovirus, and other coronaviruses. Therefore, the specificity of anti-MDA5 Ab in COVID-19 need to be further investigated. Secondly, due to limited sample size and endpoint events, we were not able to further evaluate whether anti-MDA5 Ab is an independent predictive factor for the death in COVID-19 or could be included in a risk stratification model. Thirdly, all patients were from China and it is not clear whether patients with other genetic backgrounds would have same results. Our findings are to be validated in a larger population of different ethnicities in future.

## Conclusions

Overall, we, for the first time, revealed that anti-MDA5 Ab is present in patients with COVID-19 and correlates with severe disease and poor outcomes. Early screening and serially monitoring of anti-MDA5 Ab titer has the potential to predict the disease progression of COVID-19.

## Data Availability

In this retrospective study of adult inpatients in three hospitals in China, we identified anti-MDA5 Ab in 48.2% of patients with COVID-19. The anti-MDA5 Ab positive patients tended to represent with severe disease (88.6% vs 66.9%, P<0.0001), and the titer of anti-MDA5 Ab was significantly elevated in the non-survivals (P=0.030) than that in the survivals. We also demonstrated that early profiling of anti-MDA5 Ab could distinguish severe patients from those with non-severe ones. Overall, our data suggest that early screening and serially monitoring of anti-MDA5 Ab might help identify severe COVID-19 patients with poor outcomes, or those who would probably benefit from anti-inflammation and immunosuppressive therapy. The findings are yet to be validated in a larger population of different ethnicities in future.

## Acknowledgements

We thank Dr. Jinmin Peng and Jingjing Bai for providing assistance with statistical analysis and preparing samples of anti-MDA5 Ab-related DM. We are grateful to clinicians of Wuhan Jinyintan Hospital for sample collection and Dr. colleagues of Hubei Provincial Center for Disease Control and Prevention for sample transportation.

## Contributors

CL, QW, YW, GW, LW, and HC contributed equally to this paper. CL, QW, BC and JW conceived and designed the study. CL, QW, GW, YW, LW, and HC contributed to data collection, data analysis, and data interpretation. CL, GW, and TJ performed the experiments. CH, XL, LG, LR, ML, and XZ contributed to literature search and data collection. CL, QW, BC, and JW drafted the manuscript. All authors approved the final version of the manuscript. The corresponding author attests that all listed authors meet authorship criteria and that no others meeting the criteria have been omitted.

## Finding

National Key R&D Program of China (2020YFA0707600) and CAMS Innovation Fund for Medical Sciences (2018-I2M-1-003, 2019-I2M-1-006, 2019-I2M-2-008), National Science Grant for Distinguished Young Scholars (81425001/H0104), The Beijing Science and Technology Project (D151100002115004)

## Ethical approval

The study was approved by the Research Ethics Committee of the participating hospitals and the ethical board of the Institute of Pathogen Biology, Chinese Academy of Medical Sciences (). Written informed consent was obtained from all participants before enrolment.

## Data sharing

No additional data available.

## References

1. WHO Coronavirus disease (COVID-19) Situation Report – 173. https://www.whoint/docs/default-source/coronaviruse/situation-reports/20200711-covid-19-sitrep-173Ddf?sfvrsn=949920b42

2. Wu JT, Leung K, Bushman M, et al. Estimating clinical severity of COVID-19 from the transmission dynamics in Wuhan, China. Nat Med 2020;26(4):506–10. doi: 10.1038/s41591-020-0822-7

3. Xu J, Yang X, Yang L, et al. Clinical course and predictors of 60-day mortality in 239 critically ill patients with COVID-19: a multicenter retrospective study from Wuhan, China. Crit Care 2020;24(1):394. doi: 10.1186/s13054-020-03098-9

4. Mehta P, McAuley DF, Brown M, et al. COVID-19: consider cytokine storm syndromes and immunosuppression. Lancet (London, England) 26 2020;395(10229): 1033-34. doi: 10.1016/s0140-6736(20)30628-0 [published Online First: 2020/03/21]

5. Gazzaruso C, Carlo Stella N, Mariani G, et al. High prevalence of antinuclear antibodies and lupus anticoagulant in patients hospitalized for SARS-CoV2 pneumonia. Clin Rheumatol 2020 doi: 10.1007/s10067-020-05180-7

6. Caso F, Costa L, Ruscitti P, et al. Could Sars-coronavirus-2 trigger autoimmune and/or autoinflammatory mechanisms in genetically predisposed subjects? Autoimmun Rev 2020;19(5):102524. doi: 10.1016/j.autrev.2020.102524

7. Xu Q. MDA5 should be detected in severe COVID-19 patients. Medical hypotheses 2020;143:109890. doi: 10.1016/j.mehy.2020.109890 [published Online First: 2020/06/03]

8. Giannini M, Ohana M, Nespola B, et al. Similarities between COVID-19 and anti-MDA5 syndrome: what can we learn for better care? The European respiratory journal 2020 doi: 10.1183/13993003.01618-2020

9. Zhao Q, Fang X, Pang Z, et al. COVID-19 and cutaneous manifestations: A systematic review. J Eur Acad Dermatol Venereol 2020 doi: 10.1111/jdv.16778

10. Freeman EE, McMahon DE, Lipoff JB, et al. The spectrum of COVID-19-associated dermatologic manifestations: an international registry of 716 patients from 31 countries. J Am Acad Dermatol 2020 doi: 10.1016/jjaad.2020.06.1016

11. Beydon M, Chevalier K, Al Tabaa O, et al. Myositis as a manifestation of SARS-CoV-2. Ann Rheum Dis 2020 doi: 10.1136/annrheumdis-2020-217573

12. Huang C, Wang Y, Li X, et al. Clinical features of patients infected with 2019 novel coronavirus in Wuhan, China. Lancet 2020;395(10223):497–506. doi: 10.1016/S0140-6736(20)30183-5

13. Zhao W, Zhong Z, Xie X, et al. Relation Between Chest CT Findings and Clinical Conditions of Coronavirus Disease (COVID-19) Pneumonia: A Multicenter Study. AJR Am J Roentgenol 2020;214(5):1072–77. doi: 10.2214/AJR.20.22976

14. Kawasumi H, Gono T, Kawaguchi Y, et al. IL-6, IL-8, and IL-10 are associated with hyperferritinemia in rapidly progressive interstitial lung disease with polymyositis/dermatomyositis. BioMed research international 2014;2014:815245. doi: 10.1155/2014/815245 [published Online First: 2014/05/07]

15. Sadler AJ. The role of MDA5 in the development of autoimmune disease. J Leukoc Biol 2018;103(2):185–92. doi: 10.1189/jlb.4MR0617-223R

16. Wilk AJ, Rustagi A, Zhao NQ, et al. A single-cell atlas of the peripheral immune response in patients with severe COVID-19. Nat Med 2020 doi: 10.1038/s41591-020-0944-y

17. Zhou Z, Ren L, Zhang L, et al. Heightened Innate Immune Responses in the Respiratory Tract of COVID-19 Patients. Cell Host Microbe 2020;27(6):883–90 e2. doi: 10.1016/j.chom.2020.04.017

18. Molineros JE, Maiti AK, Sun C, et al. Admixture mapping in lupus identifies multiple functional variants within IFIH1 associated with apoptosis, inflammation, and autoantibody production. PLoS Genet 2013;9(2):e1003222. doi: 10.1371/journal.pgen.1003222

19. Robinson T, Kariuki SN, Franek BS, et al. Autoimmune disease risk variant of IFIH1 is associated with increased sensitivity to IFN-alpha and serologic autoimmunity in lupus patients. J Immunol 2011;187(3):1298–303. doi: 10.4049/jimmunol.1100857

20. Martinez A, Santiago JL, Cenit MC, et al. IFIH1-GCA-KCNH7 locus: influence on multiple sclerosis risk. Eur J Hum Genet 2008;16(7):861–4. doi: 10.1038/ejhg.2008.16

21. Nejentsev S, Walker N, Riches D, et al. Rare variants of IFIH1, a gene implicated in antiviral responses, protect against type 1 diabetes. Science 2009;324(5925):387–9. doi: 10.1126/science.1167728

22. Smyth DJ, Cooper JD, Bailey R, et al. A genome-wide association study of nonsynonymous SNPs identifies a type 1 diabetes locus in the interferon-induced helicase (IFIH1) region. Nat Genet 2006;38(6):617–9. doi: 10.1038/ng1800

23. Protocol for Prevention and Control of COVID-19 (trial Edition 7). http://www.chinacdccn/jkzt/crb/zl/szkb11803/jszl11815/202003/W020200305456621460977pdf

24. Bohan A, Peter JB. Polymyositis and dermatomyositis (first of two parts). The New England journal of medicine 1975;292(7):344–7. doi: 10.1056/NEJM197502132920706

25. Bohan A, Peter JB. Polymyositis and dermatomyositis (second of two parts). The New England journal of medicine 1975;292(8):403–7. doi: 10.1056/NEJM197502202920807

26. Du Y, Liu Z, You L, et al. Pancreatic Cancer Progression Relies upon Mutant p53-Induced Oncogenic Signaling Mediated by NOP14. Cancer Res 2017;77(10):2661–73. doi: 10.1158/0008-5472.CAN-16-2339

27. Kindler E, Thiel V, Weber F. Interaction of SARS and MERS Coronaviruses with the Antiviral Interferon Response. Adv Virus Res 2016;96:219-43. doi: 10.1016/bs.aivir.2016.08.006

28. Thiel V, Weber F. Interferon and cytokine responses to SARS-coronavirus infection. Cytokine Growth Factor Rev 2008;19(2):121–32. doi: 10.1016/j.cytogfr.2008.01.001

29. Sato S, Kuwana M, Fujita T, et al. Anti-CADM-140/MDA5 autoantibody titer correlates with disease activity and predicts disease outcome in patients with dermatomyositis and rapidly progressive interstitial lung disease. Mod Rheumatol 2013;23(3):496–502. doi: 10.1007/s10165-012-0663-4

30. Zhou F, Yu T, Du R, et al. Clinical course and risk factors for mortality of adult inpatients with COVID-19 in Wuhan, China: a retrospective cohort study. Lancet (London, England) 2020;395(10229):1054–62. doi: 10.1016/s0140-6736(20)30566-3 [published Online First: 2020/03/15]

31. Wu C, Chen X, Cai Y, et al. Risk Factors Associated With Acute Respiratory Distress Syndrome and Death in Patients With Coronavirus Disease 2019 Pneumonia in Wuhan, China. JAMA Internal Medicine 2020 doi: 10.1001/jamainternmed.2020.0994

32. Liu J, Li S, Liu J, et al. Longitudinal characteristics of lymphocyte responses and cytokine profiles in the peripheral blood of SARS-CoV-2 infected patients. EBioMedicine 2020;55:102763. doi: 10.1016/j.ebiom.2020.102763

33. Ruan Q, Yang K, Wang W, et al. Clinical predictors of mortality due to COVID-19 based on an analysis of data of 150 patients from Wuhan, China. Intensive Care Med 2020 doi: 10.1007/s00134-020-05991-x

34. Zhou F, Yu T, Du R, et al. Clinical course and risk factors for mortality of adult inpatients with COVID-19 in Wuhan, China: a retrospective cohort study. The Lancet 2020 doi: 10.1016/s0140-6736(20)30566-3

35. Xu X, Han M, Li T, et al. Effective treatment of severe COVID-19 patients with tocilizumab. Proc Natl Acad Sci U S A 2020;117(20):10970–75. doi: 10.1073/pnas.2005615117

36. Cao Y, Wei J, Zou L, et al. Ruxolitinib in treatment of severe coronavirus disease 2019 (COVID-19): A multicenter, single-blind, randomized controlled trial. J Allergy Clin Immunol 2020;146(1):137–46 e3. doi: 10.1016/j.jaci.2020.05.019

37. Carbajo-Lozoya J, Muller MA, Kallies S, et al. Replication of human coronaviruses SARS-CoV, HCoV-NL63 and HCoV-229E is inhibited by the drug FK506. Virus Res 2012;165(1):112–7. doi: 10.1016/j.virusres.2012.02.002

38. Ledford H. Coronavirus breakthrough: dexamethasone is first drug shown to save lives. Nature 2020;582(7813):469. doi: 10.1038/d41586-020-01824-5

